# Strong correlations between the binding antibodies against wild type and neutralizing antibodies against omicron BA.1 and BA.2 variants of SARS-CoV-2 in individuals following booster (third dose) vaccination

**DOI:** 10.1101/2022.06.27.22276959

**Authors:** Nungruthai Suntronwong, Suvichada Assawakosri, Sitthichai Kanokudom, Ritthideach Yorsaeng, Chompoonut Auphimai, Thanunrat Thongmee, Preeyaporn Vichaiwattana, Thaneeya Duangchinda, Warangkana Chantima, Pattarakul Pakchotanon, Jira Chansaenroj, Pornjarim Nilyanimit, Donchida Srimuan, Thaksaporn Thatsanatorn, Natthinee Sudhinaraset, Nasamon Wanlapakorn, Juthathip Mongkolsapaya, Yong Poovorawan

**Affiliations:** Center of Excellence in Clinical Virology, Faculty of Medicine, Chulalongkorn University, Bangkok 10330, Thailand; Molecular Biology of Dengue and Flaviviruses Research Team, National Center for Genetic Engineering and Biotechnology (BIOTEC), National Science and Development Agency, NSTDA, Pathum Thani 12120, Thailand; Division of Dengue Hemorrhagic Fever Research, Faculty of Medicine Siriraj Hospital, Mahidol University, Bangkok 10700, Thailand; Siriraj Center of Research Excellence in Dengue and Emerging Pathogens, Faculty of Medicine Siriraj Hospital, Mahidol University, Bangkok 10700, Thailand; Wellcome Centre for Human Genetics, Nuffield Department of Medicine, University of Oxford, Oxford, OX3 7BN, UK; Chinese Academy of Medical Science (CAMS) Oxford Institute (COI), University of Oxford, Oxford, UK; The Royal Society of Thailand (FRS(T)), Sanam Sueapa, Dusit, Bangkok 10330, Thailand

**Author notes:** Equally contributed (Nungruthai Suntronwong, Suvichada Assawakosri).

**Keywords:** COVID-19, SARS-CoV-2, neutralization, omicron, antibody, correlation

## Abstract

This study examined the neutralizing activity and receptor binding domain (RBD) antibody levels against wild-type and omicron BA.1 and BA.2 variants in individuals who received three doses of COVID-19 vaccination. The relationship between the SARS-CoV-2 RBD antibody against wild-type and live virus neutralizing antibody titers against omicron BA.1 and BA.2 variants was examined. In total, 310 sera samples from individuals after booster vaccination (third dose) vaccination were tested for specific IgG wild-type SARS-CoV-2 RBD and the omicron BA.1 surrogate virus neutralization test (sVNT). The live virus neutralization assay against omicron BA.1 and BA.2 was performed using the foci-reduction neutralization test (FRNT50). The anti-RBD IgG strongly correlated with FRNT50 titers against BA.1 and BA.2. Non-linear regression showed that anti-RBD IgG with ≥148 BAU/mL and ≥138 BAU/mL were related to detectable FRNT50 titers (≥1:20) against BA.1 and BA.2, respectively. A moderate correlation was observed between the sVNT and FRNT50 titers. At detectable FRNT50 titers (≥1:20), the predicted sVNT for BA.1 and BA.2 were ≥10.57% and ≥11.52%, respectively. The study identified anti-RBD IgG and sVNT levels that predict detectable neutralizing antibodies against omicron variants. Assessment and monitoring of protective immunity support vaccine policies and will help identify optimal timing for booster vaccination.

## 1. Introduction

Detection of neutralizing antibodies helps to predict humoral immunity protection and monitor waning immunity and vaccine immunogenicity. Numerous studies have shown that a high level of neutralizing antibodies is correlated with SARS-CoV-2 protection and reduces the severity of the disease [1-3]. However, the current gold standard neutralization assay (live virus neutralization assay) has been limited to widespread use due to the need for specially trained personnel to handle the live SARS-CoV-2 virus and the need to work in a biosafety level 3 laboratory (BSL3) containment facility.

Although several commercial kits were available to detect the SARS-CoV-2 antibody and are currently used in hospital and clinical laboratories, the antigens tested derived from the ancestral strain because tests were developed before the SARS-CoV-2 variants emerged [4]. Furthermore, surrogate virus neutralization (sVNT) is widely performed to determine the ability to neutralize antibodies to block the interaction between the receptor binding domain (RBD) and human ACE2 receptors [5]. Several studies have shown that the commercial binding antibody assay and sVNT are well correlated with the gold standard results of the neutralization method against the ancestral strain [6-8]. Due to the spread of SARS-CoV-2 omicron variants, concerns have been raised as to whether preexisting immunity is sufficient to protect the omicron infection. However, the relationship between the anti-RBD IgG against wild-type and live virus neutralization assay against the omicron has been limited.

In this study, we applied non-linear regression analysis to predict the level of anti-RBD IgG and sVNT related to the detectable level of FRNT_50_ titers against omicron variants, including the BA.1 and BA.2 subvariants, in serum collected from individuals after receiving the COVID-19 booster (third dose) vaccination.

## 2. Materials and Methods

### 2.1 Participants and ethical considerations

Our study recruited 310 sera samples from individuals after receiving the booster (third dose) COVID-19 vaccination from previous studies [9, 10]. There were two primed cohorts for analysis. The first cohort was primed with two doses of AZD1222 and boosted with AZD1222, BNT162b2, 50 µg of mRNA-1273 or 100 µg of mRNA-1273 6 months after the first vaccination. The second cohort was primed with heterologous CoronaVac/ASD1222 and boosted with AZD1222, BNT162b2, and 100 µg mRNA-1273 approximately 4–5 months after the initial vaccination. The enrollment period was between November 2021 and January 2022. Blood samples were collected at day 0 and at days 28 and 90 post-booster. This study was performed following the Declaration of Helsinki and Good Clinical Practice principles. The study protocol was reviewed and approved by the Institutional Review Board of the Faculty of Medicine of Chulalongkorn University (IRB numbers 871/64 and 690/64). All participants signed a written consent before being enrolled.

### 2.2 Measurement anti-RBD IgG and sVNT

All sera samples were quantitatively measured for wild-type SARS-CoV-2 receptor binding domain (RBD) specific IgG (anti-RBD IgG) using the commercial assay, Abbott SARS-CoV-2 IgG II Quant assay (Abbott Diagnostics, Abbott Park, IL). Anti-RBD IgG was reported as a binding antibody unit (BAU/mL). The surrogate virus neutralization assay (sVNT) against variants of BA.1 was performed using a cPassTM SAR-CoV-2 neutralizing antibody detection kit (GenScript Biotech, Piscataway, NJ) as previously described [9, 10].

### 2.3 Foci reduction neutralization test (FRNT50)

For the live virus neutralization test, the foci reduction neutralization test (FRNT50) was performed using the live SARS-CoV-2 virus, which included the omicron BA.1 (GISAID accession number: EPI_ISL_8547017), BA.2 (accession number: EPI_ISL_11698090) subvariants as previously described [9, 10]. The FRNT50 titer ≥ 20 was considered a detectable level of neutralizing antibody if the neutralizing antibody titer was undetected (FRNT50 titer < 20), the FRNT50 was set as 10.

### 2.4 Statistical analysis

For statistical analysis, the predicted values of anti-RBD IgG and sVNT at FRNT50 titers ≥ 20 and ≥ 40 were determined using non-linear regression analysis and performed on the log10 transformed data. The Spearman’s rank correlation between anti-RBD IgG, sVNT, and FRNT50 titers was determined using SPSS v23.0 (IBM Corp, Armonk, NY). The r-square was calculated according to the non-linear equation using STATA v.17.0 software. A P-value <0.05 was considered statistically significant.

## 3. Results

### 3.1 Correlations between anti-RBD IgG to wild-type and FRNT50 titers against omicron

A total of 310 sera samples from individuals receiving different booster vaccination were tested for wild-type anti-RBD IgG and FRNT50 of BA.1 and BA.2 (Table 1). The FRNT50 titer ranged from undetectable (<1:20) to 3552 for BA.1 and undetectable to 3249 for BA.2 as examined on day 0 and on days 28 and 90. The correlation analysis indicated that anti-RBD IgG was strongly correlated with FRNT50 titers against BA.1 (Spearman R: 0.89, p<0.001) and BA.2 (Spearman R: 0.86, p<0.001) (S1 Table). Non-linear regression analysis showed the predicted anti-RBD IgG was 148 BAU/mL and 335 BAU/mL when the FRNT50 titers against omicron BA.1 were 20 (1.3 of log10 FRNT50 titers) and 40 (1.6 of log10 F FRNT50 titers), respectively (r2=0.79, P<0.001) (Fig 1a). In addition, the predicted anti-RBD IgG was approximately 138 BAU/mL and 298 BAU/mL when the FRNT50 titer to omicron BA.2 was 20 and 40, respectively (r2=0.73, P<0.001) (Fig 1b). When the cut-off value of 1:20 for the neutralization test was used for BA.1, the anti-RBD IgG cut-off of 148 BAU/mL showed 89.7% sensitivity and 81.4% specificity; whereas, for BA.2, the anti-RBD IgG cut-off of 138 BAU/mL showed 86.8% sensitivity and 82.9% specificity.

**Table 1.**
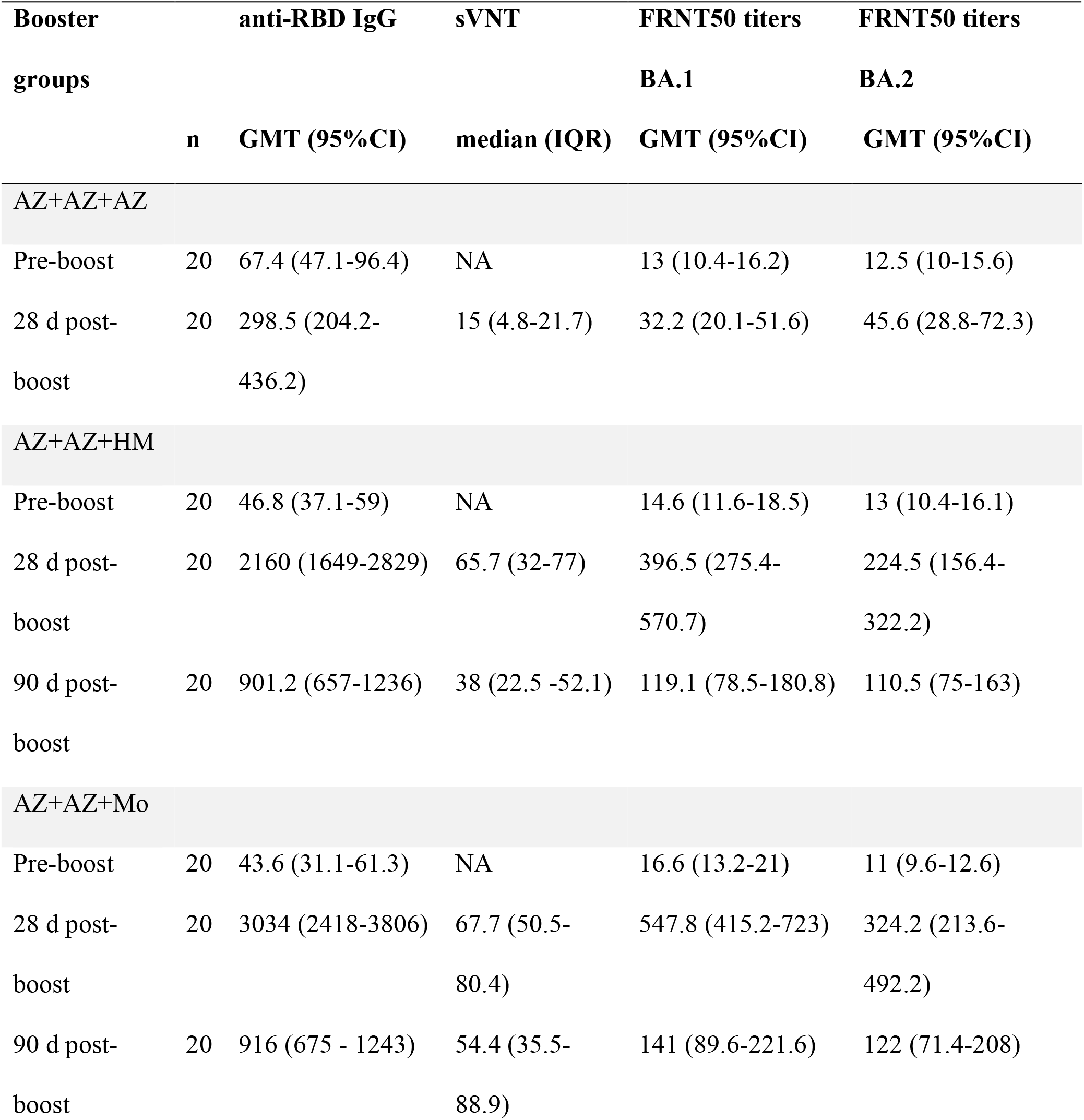
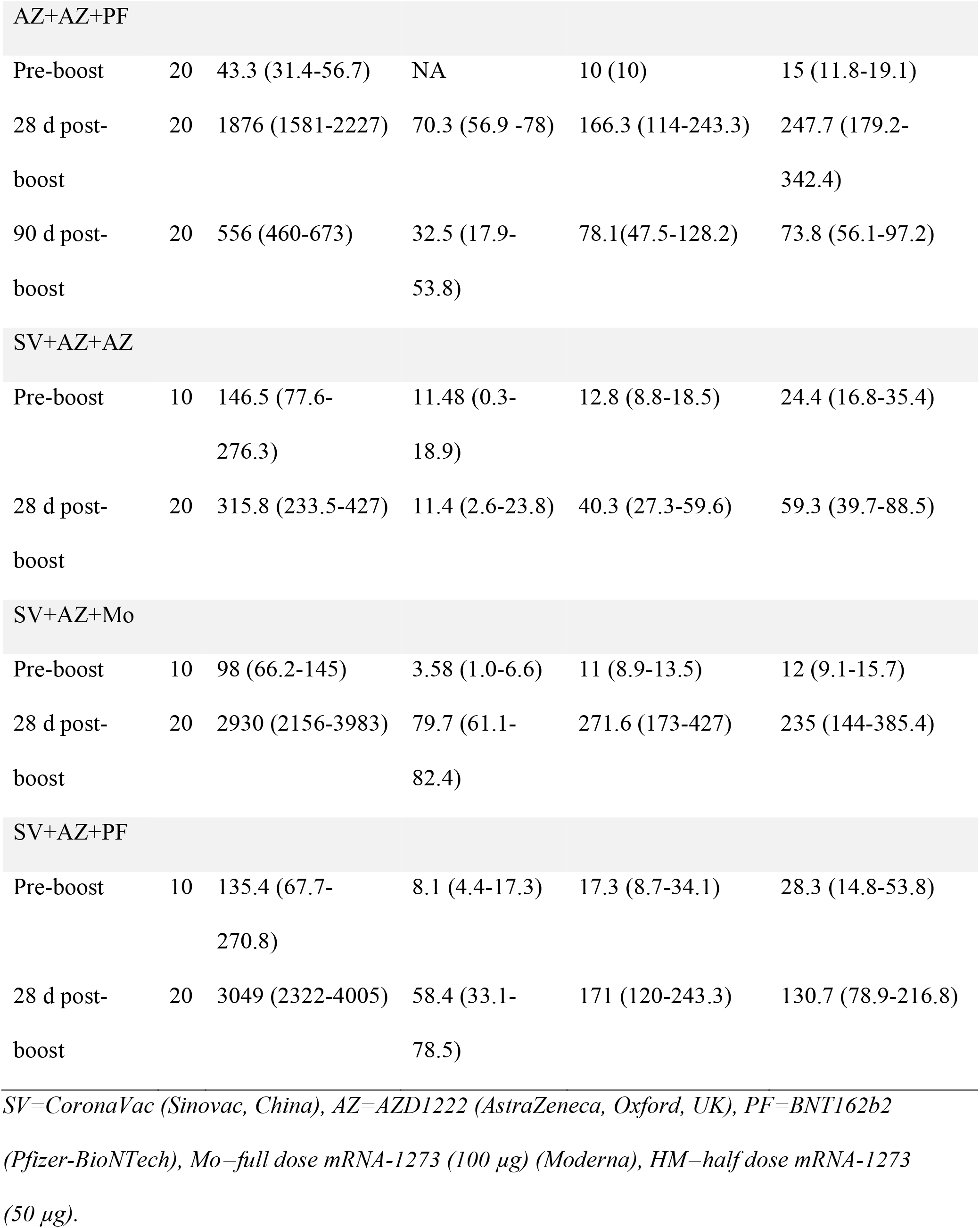
Anti-RBD IgG against wild type, sVNT against omicron, FRNT_50_ titers against BA.1 and BA.2 among the booster vaccination groups.

**Figure 1.**
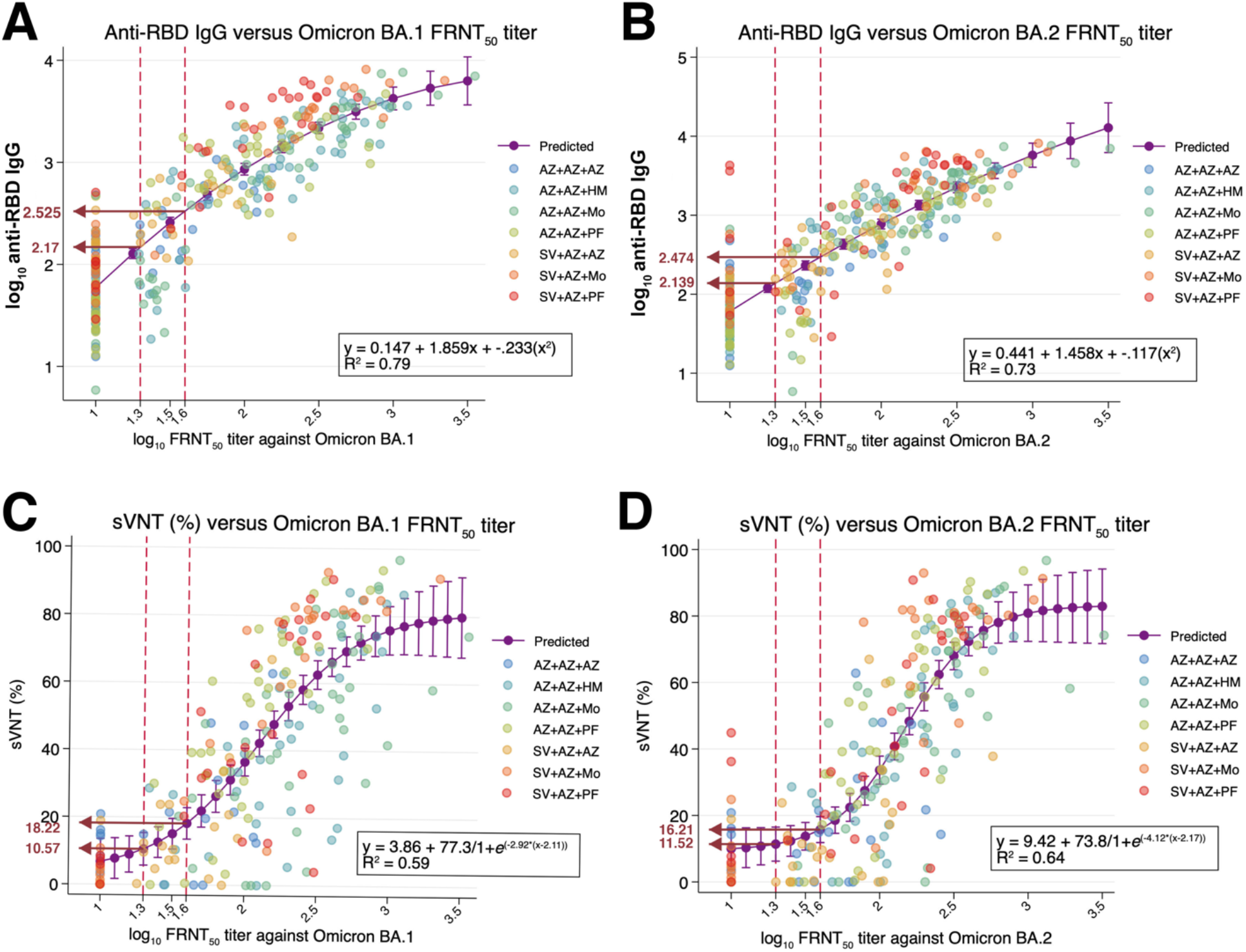
Prediction of the level of anti-RBD IgG tested against the ancestral strain and the percentage of blocking between RBD and ACE-2 interaction against omicron based on the FRNT_50_ assay against SARS-CoV-2 omicron BA.1 and BA.2 variants using non-linear regression analysis. Sera samples (n=310) from individuals with booster vaccination were tested with anti-RBD IgG, sVNT against omicron and FRNT_50_ against omicron BA.1 and BA.2. Predicted anti-RBD IgG (Panel A) and sVNT (Panel B) levels were based on the FRNT_50_ against BA.1. Predicted anti-RBD IgG (Panel C) and sVNT (Panel D) levels were based on the FRNT_50_ against BA.2. The Y axis represents log_10_ scale of anti-RBD IgG (BAU/mL). The x axis represents the log_10_ of FRNT_50_ titers. Dotted lines indicate 1.3 (FRNT_50_ titer =20) and 1.6 (FRNT_50_ titer =40). The arrows indicate the predicted level of anti-RBD IgG and percentage of inhibition from sVNT. Colored circles indicate the vaccine regimens for primary vaccine series+ booster vaccine. The r-square was calculated according to the non-linear equation using STATA v.17.0 software. SV=CoronaVac (Sinovac, China), AZ=AZD1222 (AstraZeneca, Oxford, UK), PF=BNT162b2 (Pfizer-BioNTech), Mo=full dose mRNA-1273 (100 µg) (Moderna), HM=half dose mRNA-1273 (50 µg).

### 3.2 Correlations between sVNT to omicron and FRNT50 titers against omicron

The relationship between sVNT and FRNT_50_ titers was determined and a moderate correlation between the sVNT and FRNT_50_ titers was observed (Spearman’s R=0.77 and 0.78 for BA.1 and BA.2, p<0.001) (S1 Table). Non-linear regression showed that the predicted sVNT was 10.57% and 18.22% and were related to 20 and 40 FRNT_50_ titers for BA.1 (r^2^=0.59, P<0.001). Although sVNT was 11.52% and 16.21% related to the 20 and 40 FRNT_50_0 titers for BA.2 (r^2^=0.64, P<0.001). When the cut-off level of 1:20 for the neutralization test was used for BA.1, the sVNT of ≥10.57% showed 86.4% sensitivity and 73.1% specificity, whereas for BA.2, the sVNT of ≥ 11.52% showed 82.3% sensitivity and 63.2% specificity. The ROC analysis indicated that anti-RBD IgG and sVNT provided good performance in detecting neutralizing antibodies against omicron variants (Supplementary Fig. 1a-d).

## 4. Discussion

Numerous studies have shown a strong correlation between the levels of antibody binding response, including anti-spike, anti-RBD antibodies, and neutralizing antibody titers against ancestral strain in individuals with previous COVID-19 infection or vaccination [6, 11-14]. However, there is evidence that the binding antibody was poorly correlated with neutralizing antibody titers against variants derived from B.1.1.7 and B.1.351 compared to the ancestral strain [7, 15]. In this study, we found that the anti-RBD IgG and sVNT tested by commercial kits correlated well with neutralizing antibody titers against the SARS-CoV-2 omicron variants. In addition, our data predicted anti-RBD IgG and sVNT at a detectable level of neutralizing antibodies against omicron BA.1 and BA.2 (FRNT50 titers ≥ 20). These findings suggest that boosting immunity against vaccine strain (ancestral strain) could induce cross-reactivity against omicron variants.

Anti-RBD IgG measured all antibodies targeting receptor binding sites, neutralizing antibodies, and non-neutralizing antibodies. The antigen for anti-RBD IgG detection was designed on the basis of the ancestral strain. Although more than 30 amino acid mutations were detected in the omicron variant spike protein [16], our results showed that the anti-RBD IgG and neutralizing antibody tested by FRNT50 titers against the omicron variant provided a strong correlation, which is consistent with a previous report [17]. In addition, correlations between neutralizing activity against variants of SARS-CoV-2 and RBD-specific binding antibody have been reported in samples with high binding antibody titers [7]. However, our result was inconsistent with a previous study [18], which showed that anti-RBD IgG was not correlated with the surrogate virus neutralization test against omicron variants.

In the comparison of omicron subvariants, although omicron BA.1 and BA.2 shared 12 amino acid alterations in RBD compared to wild type D614G [19], the predicted anti-RBD IgG showed higher sensitivity and specificity to detect the neutralizing antibody for omicron BA.1 than for BA.2. For sVNT, the RBD recombinant protein was designed based on BA.1 omicron variants. As expected, the sensitivity and specificity between sVNT and BA.1 were higher than BA.2.

There are several advantages to using anti-RBD IgG and sVNT to determine the antibody response against SARS-CoV-2. First, these methods do not require the live SARS-CoV-2 virus and a biosafety level 3 facility. Second, they do not require specially trained technicians and are suitable for use in hospitals and clinical laboratories. Additionally, these methods are conducted with high-throughput testing that is less time-consuming and takes 1–2 hours to complete.

There are some limitations to this study. First, the sample size was relatively small. However, we addressed the performance analysis by using samples with a wide range of antibody concentrations. Second, we did not perform the sVNT against BA.2 due to the commercial recombinant RBD protein in the production process. Furthermore, the exact level of neutralizing antibodies that protect against SARS-CoV-2 infection has not yet been established.

In conclusion, the predicted anti-RBD IgG and sVNT levels corresponding to FRNT50 titers ≥ 20 against the omicron variant showed high sensitivity and specificity. This finding underscores that anti-RBD IgG and sVNT for the omicron variants can be used to predict the presence of neutralizing antibodies against omicron BA.1 and BA.2 subvariants.

## Data Availability

All data are provided in the manuscript and supplementary files. Additional information can be requested from the corresponding author

## Funding

This research was funded by the National Research Council of Thailand (NRCT), the Health Systems Research Institute (HSRI), the Center of Excellence in Clinical Virology of Chulalongkorn University, King Chulalongkorn Memorial Hospital, MK Restaurant Group, and the Second Century Fund Fellowship of Chulalongkorn University. Thaneeya Duangchinda was funded by the National Center for Genetic Engineering and Biotechnology, BIOTEC Platform No. P2051613.

## Institutional Review Board Statement

This study was performed following the Declaration of Helsinki and Good Clinical Practice principles. The study protocol was reviewed and approved by the Institutional Review Board of the Faculty of Medicine of Chulalongkorn University (IRB numbers 871/64 and 690/64).

## Informed Consent Statement

All participants signed a written consent before being enrolled. The study was conducted according to the Declaration of Helsinki and the principles of the Good Clinical Practice Guidelines (ICH-GCP).

## Acknowledgments

We would like to thank Prof. Stephen Kerr from the Research Affairs, Faculty of Medicine, Chulalongkorn University, for the statistical analysis. We would like to thank the staff of the Center of Excellence in Clinical Virology for helping and supporting this project. Written informed consent has been obtained from the patient(s) to publish this paper.

## Conflicts of Interest

“The authors declare no conflict of interest.”

## Notes

### Competing Interest Statement

The authors have declared no competing interest.

