## Supplementary material for "Strong correlations between the binding antibodies against wild type and neutralizing antibodies against omicron BA.1 and BA.2 variants of SARS-CoV-2 in individuals following booster (third dose) vaccination": Supplementary file.docx

**S1 Table** Spearman R correlation analysis between anti-RBD IgG, surrogate virus neutralization test (sVNT) and FRNT_50_ titers against omicron BA.1 and BA.2

|  | **Spearman R** | *p* **-value** |
| --- | --- | --- |
| Anti-RBD IgG vs FRNT_50_ titers against omicron BA.1 | 0.89 | *p*<0.001 |
| Anti-RBD IgG vs FRNT_50_ titers against omicron BA.2 | 0.86 | *p* <0.001 |
| sVNT vs FRNT_50_ titers against omicron BA.1 | 0.77 | *p* <0.001 |
| sVNT vs FRNT_50_ titers against omicron BA.2 | 0.78 | *p* <0.001 |

**Figure S1** ROC analysis between anti-RBD IgG, surrogate virus neutralization test (sVNT) and FRNT_50_ titers against omicron BA.1 and BA.2

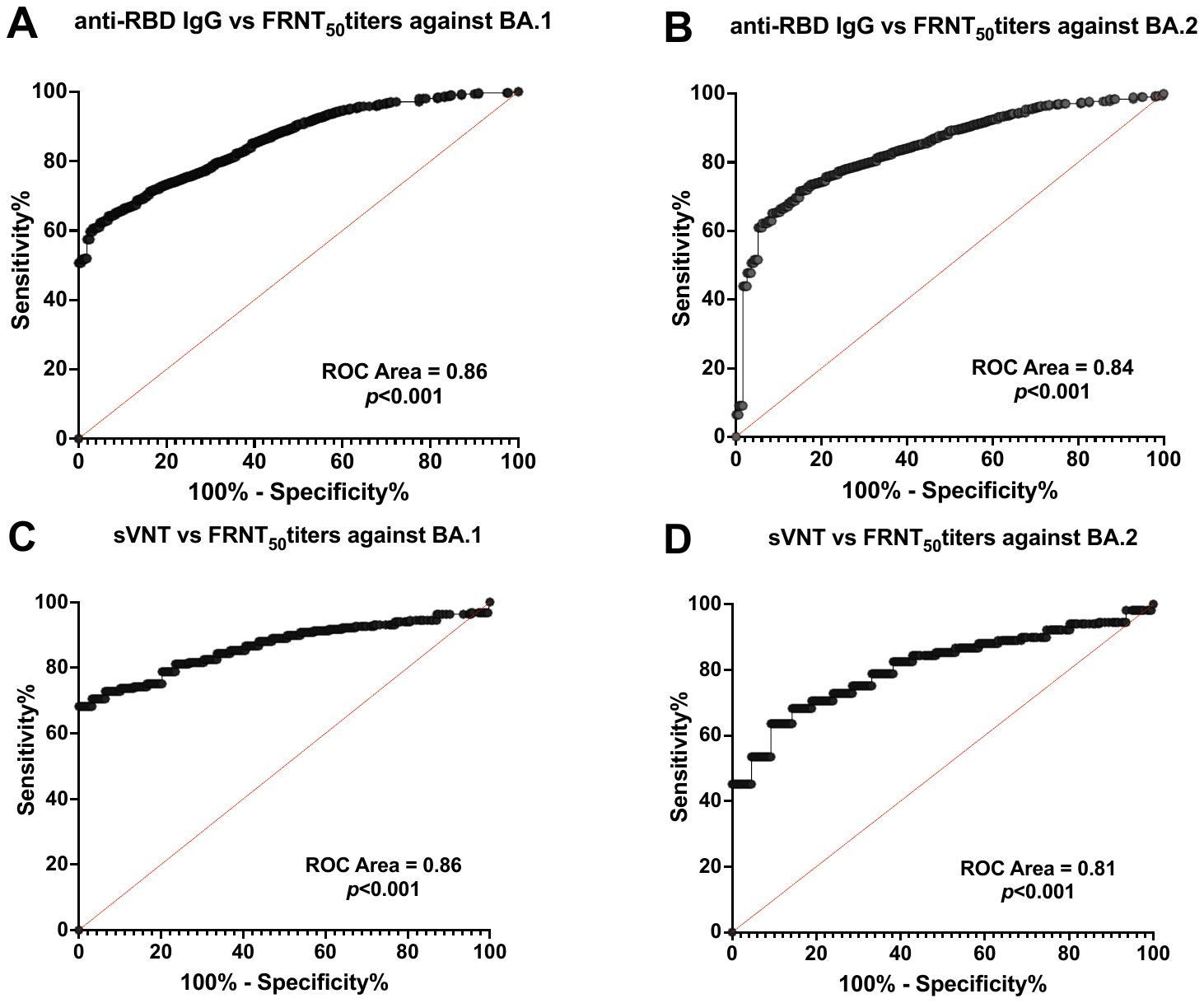
